# Measuring positive health using the My Positive Health (MPH) and Individual Recovery Outcomes Counter (I.ROC) dialogue tools: a panel study on measurement properties in a representative general Dutch population

**DOI:** 10.1101/2024.02.21.24301090

**Authors:** Vera P. van Druten, Margot J. Metz, Jolanda J.P. Mathijssen, Dike van de Mheen, Marja van Vliet, Bridey Rudd, Esther de Vries, Lenny M.W. Nahar– van Venrooij

**Author notes:** **Corresponding author** van Druten, Vera P. **STATEMENTS AND DECLARATIONS**. **Funding:** The study was funded by the Jeroen Bosch Hospital, ‘s-Hertogenbosch, the Netherlands. **Author contributions:** VvD conducted most of the statistical analyses and wrote the article. LN-vV, MM and EdV wrote the protocol, planned and supervised the research, statistical analyses and the writing process. JM conducted CFA together with VvD, supervised the statistical analyses and reviewed the manuscript. EdV, BR, MvV, MM, DvdM, LN-vV reviewed the manuscript. **Ethical approval:** The study was conducted in accordance with current public regulations, laws, and the principles of the Declaration of Helsinki. Informed consent was given by each participant to be included as a LISS-panel member. **Data availability:** All data from the LISS panel are anonymised and available upon request for researchers and policymakers. For more information see: https://www.lissdata.nl/access-data.

## Abstract

**Introduction:** Using the ‘positive health’ perspective has emerged in general healthcare. Conceptual similarities exist with the ‘recovery’ perspective in mental healthcare. Both concepts are multidimensional and focus on capability. The My Positive Health (MPH) and Individual Recovery Outcomes Counter (I.ROC) tools were developed for *dialogues*. These tools might be useful for *quantitively measuring* the *positive health* construct for monitoring and scientific purposes as well. We aimed to investigate this.

**Method:** An observational cross-sectional study was conducted in a representative general Dutch population (the LISS panel) to investigate factor structures and internal consistency from the 42-items MPH and 12-items I.ROC. After randomly splitting the dataset, exploratory factor analysis (EFA) and confirmatory factor analysis (CFA) were applied. Spearman correlation coefficient between both tools’ total scores was calculated.

**Results:** 2,457 participants completed the questionnaires. A six-factor structure was extracted for MPH (PH42) and a two-factor structure for I.ROC (I.ROC12). Explained variances were 68.1% and 56.1%, respectively. CFA resulted in good fit indices. Cronbach’s alphas were between 0.74 to 0.97 (PH42) and 0.73 to 0.87 (I.ROC12). Correlation between the total scores was 0.77.

**Conclusion:** Both PH42 and I.ROC12 are useful to quantitatively measure positive health aspects which can be summarised in sum scores in a general population. The dimensions found in this study and the corresponding item division differed from the dimensions of the original dialogue tools. Further research is recommended focussing on item reduction for PH42, factor structure of I.ROC and assessment of construct validity (in a general population) in more depth.

## INTRODUCTION

In 2011, Huber et al. introduced a new, dynamic perspective on health: *“health as the ability to adapt and self****-*** *manage in the face of social, physical, and emotional challenges”* (Huber et al., 2011). This new perspective differs from the original WHO definition of health (The World Health Organization (WHO), 2006). It focusses on someone’s capability instead of incapability, which means that people with disabilities or chronic diseases are no longer automatically seen as ‘not healthy’ (Huber et al., 2011; van Druten et al., 2022). Huber’s perspective laid the foundations for positive health, in which health is regarded as a broad construct and as an asset to living a meaningful life (Huber et al., 2016).

In the last decades, comparable focus shifts have taken place in mental healthcare for the recovery perspective. It was broadened including not only clinical recovery (reducing symptoms), but also social recovery (the role(s) someone fulfils in society), and personal recovery (living a satisfying and meaningful life) (Anthony, 1993; Davidson & Roe, 2007; Leamy et al., 2011). Taking a closer look into positive health and recovery concepts shows that both perspectives focus on a comparable multidimensional construct of health including the ability to adapt to changing conditions (Anthony, 1993; Davidson & Roe, 2007; Huber et al., 2011; Leamy et al., 2011).

To apply these perspectives of health in daily living and practice dialogue tools were developed. The My Positive Health (MPH) *dialogue* tool was developed to help citizens, patients and healthcare professionals to guide their conversation, to self-reflect and to encourage the person in question to make personalised health improvements. MPH consists of 42 items about health based on information emerged from a large study including interviews about what health is to citizens, patients and other stakeholders (Huber et al., 2016). It visually presents health in six dimensions: bodily functions, mental wellbeing, meaningfulness, quality of life, social and societal participation and daily functioning (Huber et al., 2016; Institute for positive health, n.d.). For recovery, the Individual Recovery Outcomes Counter (I.ROC) *dialogue* tool was developed, specifically for mental health related consultations (Ion et al., 2013; Monger et al., 2013). I.ROC also aims to support the conversation about broad health, self-reflection, and to make personalised recovery improvements (Rudd et al., 2020). The I.ROC consists of 12 questions visually presented in 4 domains; home, opportunity, people and empowerment (Ion et al., 2013; Monger et al., 2013). Looking at the underlying aspects representing health in the MPH and I.ROC dialogue tools it can be concluded that the content of these dialogue tools is also almost the same. Both are used as self-reported questionnaires in preparation for, or as guide during, the conversation with healthcare professionals.

To facilitate the evaluation of working with this positive health approach comprehensible quantitative outcome measures are needed. The MPH dialogue tool and its dimensions were not developed to *measure* positive health but to facilitate the conversation about positive health (Huber et al., 2016). Earlier research about measuring positive health revealed concerns about relevance, comprehensiveness and comprehensibility of the positive health model (Prinsen & Terwee, 2019). Subsequently, a shortened measurement scale, consisting of 17 out of the 42 items of the dialogue tool (PH17) was developed (Van Vliet et al., 2021). Although fit indices were adequate (Doornenbal et al., 2021), caution should be taken when generalising these results, because the original study of van Vliet et al. (Van Vliet et al., 2021) in which the PH17 was developed was based on research among citizens in just one part of the Netherlands, an eastern Dutch region, and the response rate was very low (25%) (Doornenbal et al., 2021; Van Vliet et al., 2021). In contrast to the MPH, the I.ROC is already further developed for measuring purposes. Studies for the I.ROC as measurement tool among different mental healthcare populations showed good internal consistency, test-retest reliability and concurrent validity, but with differing factor structures (Beckers et al., 2022; Dickens et al., 2017; Monger et al., 2013; Roze et al., 2020; Rudd, 2018; Rudd et al., 2020; Sportel et al., 2023). However, neither for MPH or I.ROC research at measurement properties has been conducted in a representative general population as yet.

Because of the similarities between both tools, it was hypothesised that they might both serve as a measurement tool for positive health. As a first step to develop such a generic measurement tool for positive health this study assessed measurement properties including factor structures and internal consistencies of both the MPH and I.ROC dialogue tools among a large Dutch cohort representative for a general population. It was aimed to identify if these dialogue tools measure multidimensional aspects of positive health which can be summarised in sum scores. Additionally, coherence between the MPH and the I.ROC was assessed.

## METHODS

### Study design

An observational cross-sectional research design was used to evaluate factor structures and internal consistency of the MPH and I.ROC. Correlation between the MPH and the I.ROC dialogue tools was calculated in a representative cohort from the general Dutch population, the Longitudinal Internet Studies for the Social sciences (LISS) panel.

### Participants

The LISS panel (http://www.lissdata.nl) is a non-commercial online longitudinal research panel, operational since 2007 and includes a representative sample of approximately 7,000 individuals from approximately 5,000 households from the Dutch general population. The panel includes both healthy people and people with health conditions, all living at home. The panel was carefully composed based on a true probability distribution drawn from the population register by Statistics Netherlands (Scherpenzeel & Das, 2010). To become a LISS panel member, at least one person in the household should master the Dutch language. To minimise selection bias, households were provided with a computer and internet connection if they could not otherwise participate.

### Data collection

LISS panel members complete monthly online questionnaires and are paid for each completed questionnaire. Response rates for this panel are high. For example, 5,714 (83.6%) LISS panel members completed a LISS core study module about health in November 2020 (https://www.lissdata.nl/research/liss-core-study). For this study, the selected study population was asked to additionally fill out the MPH and I.ROC dialogue tool questionnaires. Rule of thumb for sample size calculation is four to ten respondents per item of a questionnaire with a minimum of 100 patients according to de Vet et al. (2018). A study population of 2,500 respondents is adequate to assess factor structures of the data and their goodness of fit in two separate groups (n=1250). To achieve the required number (n=2500) counting on a response rate of 75%, 3,246 adults (≥18 years, one per household) of the LISS panel were randomly selected.

### Dialogue tools MPH and I.ROC

The MPH dialogue tool consists of 42 statements scored on an 11-point scale ranging from 0 ‘totally disagree’ to 10 ‘totally agree’. The I.ROC dialogue tool consists of 12 items on a six-point scale from 1 ‘never’ to 6 ‘all the time’. Higher scores represent better health (see Supplementary tables 1 and 2). In this study both dialogue tools are used as self-reported questionnaires.

### Evaluation of measurement properties/analytical plan

First, exploratory factor analysis (EFA) was used to explore factor structures and generate sum scores per factor. For this purpose, the extraction method principal component analysis (PCA) was applied. PCA (also called exploratory factor analysis) is a statistical technique used for analysing the interrelations among a set of variables (or questions in the questionnaire) and for explaining these interrelations in terms of a reduced number of variables, called factors (de Vet et al., 2018; Floyd & Widaman, 1995; Velicer & Jackson, 1990). Next, to assess if the extracted factor structure also holds in another group, confirmatory factor analysis (CFA) was applied in another part of the sample. The dataset was randomly split; group 1 for EFA and group 2 for CFA. Last, per factor internal consistency was explored, and coherence between the total scores of both tools was assessed. Statistical analyses were performed using software package IBM SPSS version 27, except for CFA which was analysed with IBM SPSS Amos version 27.

### Exploratory factor analysis

Step 1: associations between all items were assessed with Kaiser-Meyer-Olkin (KMO) and Bartlett’s test correlation in data group 1; these should be >0.5 with significance level <0.05 to have sufficient correlations for factor analysis. Items that do not correlate with any other item (<0.2) were discussed for its content. Item correlations of >0.9 might measure the same thing. Items with low or high correlations (<0.2 and >0.9) could be retained or deleted, depending on the qualitive content discussion about the importance for the construct to be measured.

Step 2: the number of factors was identified with Oblimin with Kaiser Normalisation rotation, because items and factors were not expected to be independent from each other. The number of factors was determined with a criterion eigenvalue >1 and inspection of the scree plot for confirmation. Highest factor loadings of items were used as guidance to divide the items over the factors with a minimum threshold of 0.32 (21). Factor loadings of >0.71 are considered excellent, >0.63 very good, >0.55 good and >0.45 fair (Tabachnick & Fidell, 2007). Since our aim was not to reduce items but to assess structures and to generate sum scores, we chose the relatively low cut-off point for factor loadings of >0.32 for including items (Tabachnick & Fidell, 2007).

Step 3: item content and factor structure were discussed by the research team. The items were assigned to the factors on which they loaded the highest, unless an item conceptually fitted better in another factor according to the research team; then they were relocated.

Step 4: inter-item correlations per factor were assessed; values should be between 0.2 and 0.5. Inter-item correlations greater than 0.7 suggest that they may measure the same thing (de Vet et al., 2018).

#### Confirmatory factor analysis

Step 5: Extracted factor structures were assessed for goodness of fit using CFA. Maximum likelihood (ML) was used as estimation method. Goodness of fit indices included chi-square (X^2^). A non-significant X^2^ is desirable, however in a large sample, the X^2^ is usually significant. Comparative fit index (CFI) and Root mean square error of approximation (RMSEA) were therefore also used. CFI values between 0.90 and 0.95 are indicators of acceptable fit and >0.95 indicates superior model fit (Byrne, 2010; Hu & Bentler, 1999). RMSEA values <0.05 represent good fit, 0.05-0.08 acceptable fit, >0.08 medium fit and >0.1 poor fit (Byrne, 2010; Hu & Bentler, 1999).

#### Internal consistency

Step 6: Cronbach’s alphas were calculated to further explore internal consistency; values of factors between 0.7 and 0.9 with a minimum of 3 items were considered good (de Vet et al., 2018).

#### Total scores and sum scores per factor

Step 7: To describe the distribution of the data, mean, median, standard deviation, minimum, maximum, skewness and kurtosis (< -1 and < 1) of the factor scores and total scores were calculated. If ≥ 15% of the respondents scored lowest possible scores this suggests that floor effects exists (de Vet et al., 2018). If ≥ 15% of the respondents scored highest possible scores this suggests ceiling effects (de Vet et al., 2018).

#### Correlation between total scores

Step 8: Correlation between the total scores of MPH and I.ROC was calculated with Spearman’s correlation coefficient to assess if coherence exists between the tools. A value >0.4 suggests moderate to strong correlation (de Vet et al., 2018).

### Ethical considerations

The study was conducted in accordance with current public regulations, laws, and the principles of the Declaration of Helsinki. Ethical approval was given by the METC Brabant (Tilburg, the Netherlands, study number NW2024-15). Informed consent was given by each participant to be included as a LISS panel member and for each monthly questionnaire (Scherpenzeel & Das, 2010). The General Data Protection Regulation (GDPR) was followed. All data from the LISS panel are anonymised and open access available for researchers and policymakers upon request.

## RESULTS

### Characteristics of the respondents

The response rate was high (76%) with 777 respondents not responding. Twelve respondents did not fill out the questionnaires completely and this small number was excluded, leaving 2,457 respondents for the analyses.

Characteristics of the included respondents are shown in Table 1. The response group (n=2,457) was randomly split in group 1 for EFA (n=1,199) and group 2 for CFA (n=1,258) as described in the Methods section.

**Table 1:**
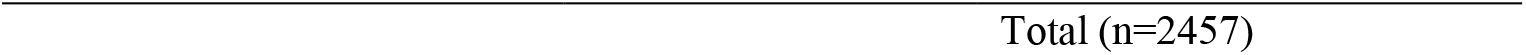
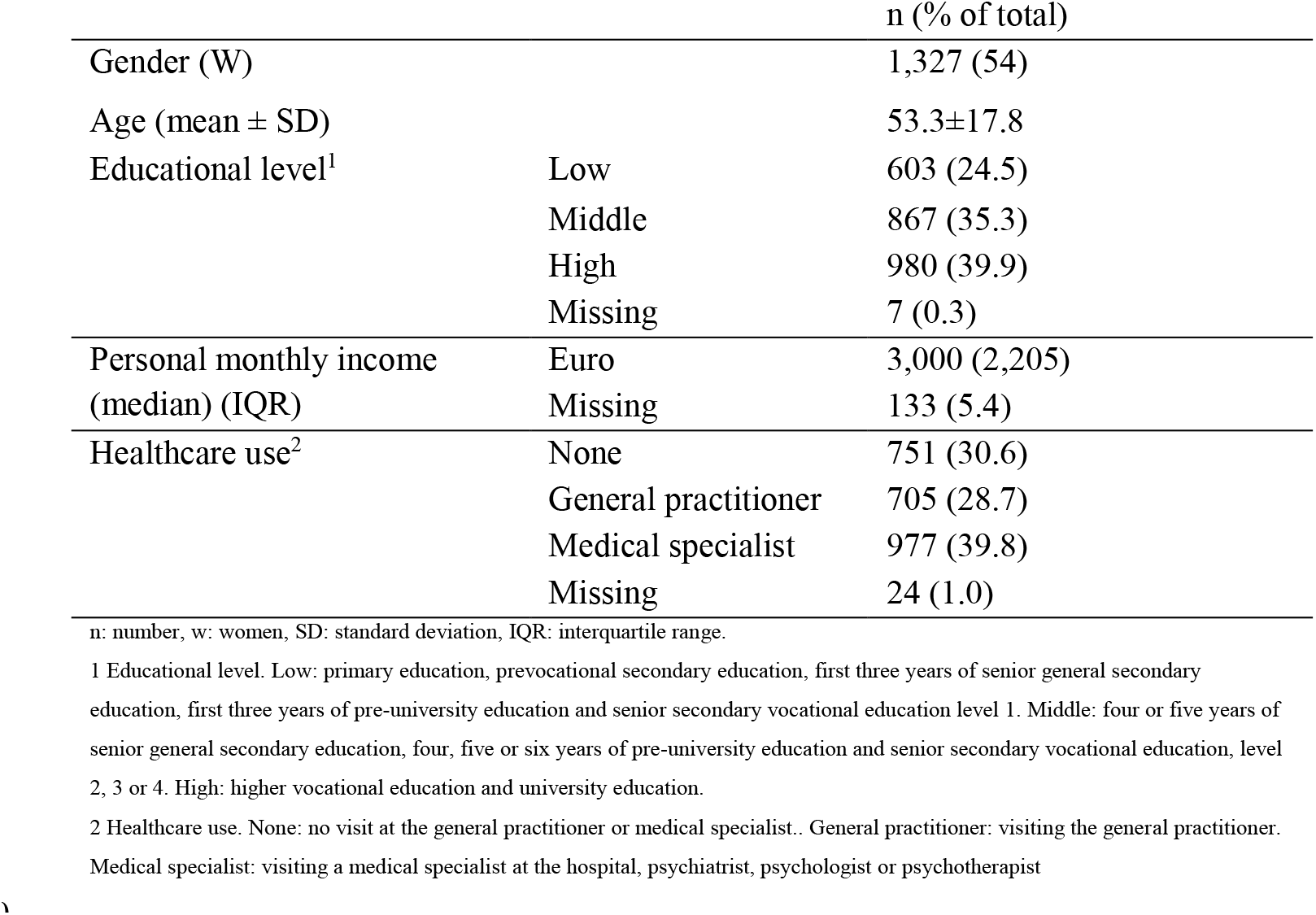
characteristics of the respondents.

|  |  | n (% of total) |
| --- | --- | --- |
| Gender (W) |  | 1,327 (54) |
| Age (mean ± SD) | | 53.3±17.8 |
| Educational level<sup>1</sup> | Low | 603 (24.5) |
|  | Middle | 867 (35.3) |
|  | High | 980 (39.9) |
|  | Missing | 7 (0.3) |
| Personal monthly income (median) (IQR) | Euro | 3,000 (2,205) |
|  | Missing | 133 (5.4) |
| Healthcare use<sup>2</sup> | None | 751 (30.6) |
|  | General practitioner | 705 (28.7) |
|  | Medical specialist | 977 (39.8) |
|  | Missing | 24 (1.0) |

n: number, w: women, SD: standard deviation, IQR: interquartile range.
1 Educational level. Low: primary education, prevocational secondary education, first three years of senior general secondary education, first three years of pre-university education and senior secondary vocational education level 1. Middle: four or five years of senior general secondary education, four, five or six years of pre-university education and senior secondary vocational education, level 2, 3 or 4. High: higher vocational education and university education.
2 Healthcare use. None: no visit at the general practitioner or medical specialist.. General practitioner: visiting the general practitioner. Medical specialist: visiting a medical specialist at the hospital, psychiatrist, psychologist or psychotherapist

### My Positive Health (MPH)

#### Exploratory factor analysis (EFA)

Step 1: KMO and Bartlett’s test was statistically significant (0.97; p <=.01). Item correlations showed that item 3 (having complaints or pain) of the MPH dimension Bodily Functions (BF) did not correlate (<0.2) with 15 items from the MPH dimensions Mental Wellbeing (MW), Meaningfulness (MF), Quality of Life (QL), Social and societal Participation (SP) and Daily Functioning (DF): MW10, MW13, MF21, QL27, QL28, SP29, SP31, SP32, SP33, SP35, DF36, DF37, DF40, DF41, DF42 (for content of these items see Table 2). Based on relevance of the content and its correlation with other items, item BF3 was not deleted.

**Table 2:** factor loadings from the exploratory factor analysis of MPH.

| Pattern Matrix MPH |  | Factor |  |  |  |  |  |
| --- | --- | --- | --- | --- | --- | --- | --- |
|  |  | Acceptance,<br>meaningfulness and<br>satisfaction with life | Physical<br>health and<br>functioning | Self-management | Social<br>network and<br>societal roles | Personal<br>development | Cognition |
| Item number<br>of the<br>dialogue tool | Item description |  |  |  |  |  |  |
| BF1 | Feeling healthy | 0.195 | **0.769** | 0.027 | -0.032 | 0.016 | 0.028 |
| BF2 | Feeling fit | 0.206 | **0.781** | 0.052 | -0.039 | -0.002 | 0.003 |
| BF3 | Having complaints or pain | -0.031 | **0.450** | -0.097 | 0.062 | -0.018 | 0.193 |
| BF4 | Sleeping pattern | 0.395 | **0.422** | -0.061 | 0.022 | -0.276 | 0.254 |
| BF5 | Eating pattern | 0.241 | **0.364** | 0.248 | 0.087 | -0.250 | 0.197 |
| BF6 | Physical condition | 0.069 | **0.783** | 0.046 | 0.021 | 0.048 | -0.013 |
| BF7 | Exercise | -0.107 | **0.877** | 0.061 | 0.049 | 0.109 | -0.118 |
| MW8 | Being able to remember things | 0.129 | 0.057 | 0.120 | 0.074 | 0.140 | **0.654** |
| MW9 | Being able to concentrate | 0.303 | 0.021 | 0.161 | 0.027 | 0.099 | **0.610** |
| MW10 | Being able to communicate | -0.297 | 0.095 | 0.250 | 0.327 | 0.157 | **0.429** |
| MW11 | Being cheerful | **0.653** | 0.159 | -0.040 | 0.154 | 0.089 | 0.102 |
| MW12 | Accepting yourself | **0.572** | 0.037 | 0.206 | 0.054 | 0.043 | 0.145 |
| MW13 | Being able to handle changes | 0.106 | -0.034 | 0.045 | 0.131 | **0.518** | 0.259 |
| MW14 | Having control | **0.534** | 0.026 | 0.201 | 0.105 | 0.163 | 0.123 |
| MF15 | Having a meaningful life | **0.573** | 0.049 | 0.045 | 0.249 | 0.183 | 0.005 |
| MF16 | Being high-spirited | **0.673** | 0.105 | 0.046 | 0.129 | 0.102 | 0.033 |
| MF17 | Wanting to achieve ideals | 0.310 | 0.161 | -0.115 | -0.008 | **0.643** | 0.051 |
| MF18 | Feeling confident about own future | **0.558** | 0.161 | 0.033 | 0.093 | 0.344 | -0.019 |
| MF19 | Accepting life | **0.645** | -0.039 | 0.165 | 0.048 | 0.135 | 0.087 |
| MF20 | Being grateful | **0.624** | 0.008 | 0.114 | 0.135 | 0.160 | 0.002 |
| MF21 | Continue learning | 0.182 | 0.118 | 0.020 | 0.002 | **0.660** | 0.017 |
| QL22 | Enjoyment | **0.676** | 0.086 | 0.021 | 0.253 | 0.060 | -0.013 |
| QL23 | Being happy | **0.699** | 0.082 | -0.004 | 0.260 | 0.034 | -0.023 |
| QL24 | Feeling good | **0.708** | 0.235 | -0.009 | 0.097 | 0.051 | 0.005 |
| QL25 | Feeling well-balanced | **0.751** | 0.064 | 0.129 | 0.053 | 0.047 | 0.036 |
| QL26 | Feeling safe | **0.393** | 0.074 | 0.185 | 0.256 | 0.028 | 0.078 |
| QL27 | Living conditions | 0.335 | -0.044 | 0.224 | **0.423** | -0.140 | -0.017 |
| QL28 | Having enough money | 0.219 | 0.035 | **0.598** | 0.147 | -0.121 | -0.231 |
| SP29 | Social contacts | 0.111 | 0.013 | -0.038 | **0.786** | 0.013 | 0.051 |
| SP30 | Being taken seriously | -0.023 | -0.021 | 0.047 | **0.792** | 0.081 | 0.092 |
| SP31 | Doing fun things together | 0.043 | 0.070 | -0.070 | **0.899** | -0.014 | -0.043 |
| SP32 | Having the support of others | -0.014 | -0.013 | -0.036 | **0.939** | -0.038 | 0.015 |
| SP33 | Belonging | 0.099 | -0.015 | 0.018 | **0.864** | -0.070 | -0.028 |
| SP34 | Doing meaningful things | 0.228 | 0.212 | 0.103 | **0.401** | 0.181 | -0.169 |
| SP35 | Being interested in society | 0.011 | -0.002 | 0.181 | **0.418** | 0.263 | -0.011 |
| DF36 | Looking after yourself | -0.171 | 0.290 | **0.633** | 0.052 | 0.092 | 0.065 |
| DF37 | Knowing your limitations | 0.001 | 0.014 | **0.754** | 0.017 | 0.011 | 0.233 |
| DF38 | Knowledge of health | -0.022 | 0.157 | **0.651** | 0.088 | 0.019 | 0.201 |
| DF39 | Managing time | 0.132 | 0.001 | **0.634** | -0.062 | 0.073 | 0.238 |
| DF40 | Managing money | 0.147 | -0.036 | **0.828** | 0.001 | -0.069 | -0.070 |
| DF41 | Being able to work | -0.089 | **0.526** | 0.291 | 0.058 | 0.266 | -0.223 |
| DF42 | Asking for help | 0.065 | -0.044 | **0.452** | 0.178 | 0.175 | -0.154 |

Extraction method Principal Component Analysis. Rotation Method: Oblimin with Kaiser Normalisation.
Items with factor loadings >0.32 are preserved. In bold: items belonging to the subscale on behalf of highest factor loading.
BF: bodily functions, MW: mental wellbeing, MF: meaningfulness, QL: quality of life, SP: social and societal participation, DF: daily functioning (Huber et al., 2016).

Step 2: the extraction method PCA and Oblimin rotation revealed a six-factor structure with an eigenvalue >1 explaining 68.1% of the variance. Factor loadings ranged from 0.36 to 0.94. After careful consideration within the research team, the factors were named: ‘Acceptance, meaningfulness and satisfaction with life’, ‘Physical health and functioning’, ‘Self-management’, ‘Social network and societal roles’, ‘Personal development’ and ‘Cognition’. For details of factors, items and factor loadings see Table 2.

Step 3: the research team discussed item distribution among the factors on its content; no changes were made based on the discussion.

Step 4: inter-item correlations showed that 48.1% of the correlations within the factor ‘Acceptance, meaningfulness and satisfaction with life’, 14.3% within the factor ‘Physical health and functioning’, 4.8% within the factor ‘Self-management’, 25% within the factor ‘Social network and societal roles’, none within the factor ‘Personal development’ and 33.3% within the factor ‘Cognition were >0.7 (For more details see Supplementary tables 3 to 8.).

#### Confirmatory factor analysis

Step 5: the extracted six-factor structure had an acceptable fit in CFA with significant X^2^ (5290.328; degrees of freedom 790; *p* <= 0.01), CFI of 0.90 and RMSEA of 0.067 with a 90% confidence interval of 0.066 to 0.069.

#### Internal consistency of MPH

Step 6: Cronbach’s alphas of the extracted factors ranged from 0.74 to 0.97; 0.969 (Acceptance, meaningfulness and satisfaction with life), 0.866 (Physical health and functioning), 0.860 (Self-management), 0.923 (Social network and societal roles), 0.742 (Personal development) and 0.801 (Cognition).

#### Sum scores per factor and total score of MPH

Step 7: Results are shown in Table 3. No floor or ceiling effect was present on the factors or total score; no more than 8.3% of the respondents scored the highest possible score and none of the respondents scored the lowest possible score (data not shown).

**Table 3:** Total scores and sum scores per factor of PH42 for the total response group (n=2457)

|  | Acceptance,<br>meaningfulness and<br>satisfaction with life<br>(min-max score on<br>the factor 0-130) | Physical health<br>and<br>functioning<br>(min-max<br>score on the<br>factor 0-80) | Self-management<br>(min-max<br>score on the<br>factor 0-70) | Social<br>network and<br>societal roles<br>(min-max<br>score on the<br>factor 0-80) | Personal<br>development<br>(min-max<br>score on the<br>factor 0-30) | Cognition<br>(min-max<br>score on<br>the factor<br>0-30) | Total score<br>(min-max<br>score on<br>the total<br>score 0-420) |
| --- | --- | --- | --- | --- | --- | --- | --- |
| Mean | 98.4 | 57.5 | 58.2 | 62.9 | 21.9 | 23.8 | 322.6 |
| Median | 102 | 60 | 59 | 64 | 22 | 24 | 330 |
| SD | 20.4 | 13.0 | 8.6 | 11.4 | 4.8 | 4.3 | 53.4 |
| Skewness | -0.95 | -0.7 | -1.0 | -1.02 | -0.68 | -0.91 | -0.79 |
| Kurtosis | 1.12 | 0.408 | 1.31 | 1.64 | 0.47 | 1.09 | 0.75 |
| Min | 9 | 7 | 18 | 4 | 3 | 2 | 104 |
| Max | 130 | 80 | 70 | 80 | 30 | 30 | 420 |

### Individual Recovery Outcomes Counter (I.ROC)

#### Exploratory factor analysis

Step 1: KMO and Bartlett’s test was significant (0.92; p <=.01). Item correlations showed that only item 8 (Social network) did not correlate (<0.2) with items 2 (life skills) and 3 (safety and comfort). Based on relevance of the content and its correlation with other items, item PE8 was not deleted.

Step 2: the extraction method PCA with Oblimin rotation revealed a two-factor structure with an eigenvalue >1 explaining 56.1% of the variance. Factor loadings ranged from 0.37 to 0.87. After careful consideration within the research team, the factors were named: ‘Wellbeing, control, network and meaningfulness’ and ‘Health, safety and abilities’ (Table 4).

**Table 4:** Factor loadings from the exploratory factor analysis of I.ROC.

| Pattern Matrix I.ROC |  | Factor |  |
| --- | --- | --- | --- |
|  |  | Wellbeing, control, network and meaningfulness | Health, safety and abilities |
| Item number of questionnaire | Description |  |  |
| HO1 | Mental health | **0.453** | 0.438 |
| HO2 | Life skills | -0.098 | **0.872** |
| HO3 | Safety and comfort | 0.116 | **0.677** |
| OP4 | Physical health | 0.213 | **0.633** |
| OP5 | Exercise and activity | 0.375 | **0.369** |
| OP6 | Purpose and direction | **0.577** | 0.238 |
| PE7 | Personal network | **0.671** | 0.058 |
| PE8 | Social network | **0.778** | -0.270 |
| PE9 | Valuing myself | **0.697** | 0.147 |
| EM10 | Participation and control | **0.775** | 0.026 |
| EM11 | Hope for the future | **0.730** | 0.124 |
| EM12 | Self-management | **0.675** | 0.159 |

Extraction method Principal Component Analysis. Rotation Method: Oblimin with Kaiser Normalization.
Items with factor loadings $>0.32$ are preserved. In bold: items belonging to the subscale.
HO: Home, OP: opportunity, PE: people, EM: empowerment (Ion et al., 2013; Monger et al., 2013).

Step 3: the research team concurred on item distribution for 11 items. Item 5 (exercise and activity) loaded almost equally on both factors. This item was relocated to the factor ‘Health, safety and abilities’ following discussion about the content of the I.ROC dimensions and related items. For details of factors, items and factor loadings see Table 4.

Step 4: for both factors all inter-item correlations were <0.7 (see Supplementary tables 9 and 10).

#### Confirmatory factor analysis

Step 5: the two-factor structure extracted from EFA had an acceptable fit in CFA with significant X^2^ (321.503; degrees of freedom 47; *p* <= 0.01), CFI of 0.96 and RMSEA of 0.068 with a 90% confidence interval of 0.061 to 0.075.

#### Internal consistency of I.ROC

Step 6: Cronbach’s alphas of the extracted factors were 0.87 (Wellbeing, control, network and meaningfulness) and 0.73 (Health, safety and abilities).

#### Sum scores per factor and total scores of I.ROC

Step 7: Results are shown in Table 5. No more than 9.3% of the respondents scored the highest score on one of the factors or the total score and no respondents scored the lowest score (data not shown).

**Table 5:** Total scores and sum scores per factor of I.ROC12 for the total response group (n=2457)

|  | Wellbeing, control, network and meaningfulness<br>(min-max score on the factor 8-48) | Health, safety and abilities<br>(min-max score on the factor 4-24) | Total score<br>(min-max score on the total score 12-72) |
| --- | --- | --- | --- |
| Mean | 35.5 | 20.0 | 55.4 |
| Median | 36 | 20 | 56 |
| SD | 6.5 | 3.0 | 8.8 |
| Skewness | -0.45 | -0.89 | -0.55 |
| Kurtosis | 0.11 | 0.73 | 0.27 |
| Min | 10 | 7 | 21 |
| Max | 48 | 24 | 72 |

### Correlation between total scores of MPH and I.ROC

Step 8: Spearman correlation coefficient between total scores of MPH and I.ROC was 0.77 and significant (*P<*=0.01).

## DISCUSSION

This study assessed whether the original items of the *dialogue* tools MPH and I.ROC can be used for *measuring multiple aspects of positive health summarised in sum scores* among a general (Dutch) population. For the MPH tool, factor analysis resulted in a six-factor structure including all 42 items and for the I.ROC tool in a two-factor structure including all 12 items. For both tools, CFA confirmed acceptable fit to the data. Internal consistency was good, except for two of the extracted factors of the MPH tool in which internal consistency was too high (>0.9). The total scores of the tools were highly correlated (0.77). To differentiate the original dialogue tools MPH and I.ROC from the measurement tools, these measurement tools (questionnaires) are further called the PH42 and I.ROC12. These results imply important points for consideration as described below.

It is important to realise that different methodological approaches are used to identify dimensions depending on the aim of the instrument; i.e., whether it is meant to be a dialogue tool or a quantitative measurement tool. These different aims may lead to different results. The study of Huber et al. (2016) intended to get more insight in what health meant for different stakeholder groups (patients, practitioners, citizens and policymakers) using a qualitative research design. The indicators of health that emerged from this study led to the operationalisation of the concept of positive health (Huber et al., 2016). The concept was further elaborated and an MPH dialogue tool including six dimensions was developed, aiming to support the conversation about someone’s health in daily practice from a positive health perspective as well as to support self-evaluation (Huber et al., 2016). The aim of our study was to evaluate the items of the MPH and I.ROC dialogue tools for quantitative measuring purposes among a general population by assessing their factor structure and generating factor sum scores. In addition, the aim was not to reduce items, as long as the items did not hamper a clear interpretation. Therefore, the more conservative cut-off point for factor loadings of >0.32 (Tabachnick & Fidell, 2007) instead of the >0.5 cut-off point aiming to reduce items (de Vet et al., 2018), was chosen. Compared to the MPH dialogue tool, we identified differences in the division of items over the dimensions of the investigated PH42 measurement tool. Both the MPH dialogue tool and the PH42 measurement tool correspond to the positive health construct. The results showed that the measurement tool is still multidimensional while no items (and thus aspects of positive health) were deleted. Similar issues were seen for the I.ROC dialogue tool. Initially, the four dimensions of the I.ROC dialogue tool were also developed for visual presentation and to guide the conversation (Monger et al., 2012). In the current study differences in the division of items over two dimensions of the I.ROC12 measurement tool were identified. For *quantitative measuring* purposes the dimensions retrieved from our study are the best fit.

The factor structure found for I.ROC12 in our general population did not correspond with the results from earlier research aiming to develop a measurement tool based on the I.ROC for a mental healthcare population. While our study resulted in two factors, these studies resulted in one up to three factors (Beckers et al., 2022; Dickens et al., 2017; Monger et al., 2013; Rudd et al., 2020). These differences can most likely be explained by the differences in the study populations, because perceptions and values of aspects of health may differ between a mental healthcare population and the general population (van Druten et al., 2022). Furthermore, cross-cultural measurement invariance between our Dutch population and the Scottish populations, in which the I.ROC was developed and first validated, may be present indicating that comparable groups of respondents among different cultures score differently on a questionnaire (de Vet et al., 2018). Also, while this study was being performed, the COVID-19 pandemic struck, which can have led to different scores particularly on the questions about social network and activities. However, the PH17 which was developed pre-COVID showed a factor structure comparable to the PH42 (Van Vliet et al., 2021). In contrast to the I.ROC12, for the PH42, no research into factor structures in other than a general population has been conducted so far. If the PH42 and I.ROC12 will be used in other populations, more research is needed to assess if the found factor structures for the PH42 and the I.ROC12 also hold.

Although the PH42 seems useful for measurement purposes, the results imply that future research into item reduction is advisable. Two factors ‘Acceptance, meaningfulness and satisfaction with life’ and ‘Social network and societal roles’ had high internal consistency (>0.9) meaning that a redundancy of items is present; there was also a high amount of inter-item correlations (>0.7) in these factors (48.7% and 25%), meaning that some items might measure the same thing. Furthermore, it should be stressed that PH42 includes items that loaded substantially (>0.3) on more than one factor (6/42). These items were judged not to hamper a clear interpretation of the factor and were therefore retained. These results imply added value of item reduction for the PH42 to develop a clarified, comprehensible and comprehensive measurement tool.

We conclude the PH42 to be the first choice for quantitively measuring the multidimensional concept of positive health in a general population. The I.ROC12 – although developed for a different purpose (recovery) – is a potential alternative. We found a high coherence between the total scores of the tools supporting that they measure a comparable construct. The I.ROC12 is shorter, however, the PH42 gives more information and the explained variance of the PH42 is higher. The results of our study offer some support for the use of the PH17 (Van Vliet et al., 2021) as a shorter alternative: all except one item of the dimensions of the PH17 showed overlap with the dimensions of the PH42. However, Van Vliet et al. (2021) did not apply the preferred methodological approach for item reduction as published by de Vet et al. (2018), when developing the PH17.

This approach implies item reduction based on repeated factor analyses after removal of each item including judgement of factor loadings and inter-item correlations, content discussion, and maintaining acceptable internal consistencies. As a next step in the validation process of both the PH42 and I.ROC12 in a general population, assessment of construct validity in more depth is recommended.

This study has some limitations. A minor limitation of the selection procedure was that at least one member of the household had to master the Dutch language, whereby migrants were relatively underrepresented. Also, the members of the participating households had to be able to use a computer, which may have led to some selection bias. However, this study is robust in terms of its large sample size, the high response rate and the representativeness of the general Dutch population (Scherpenzeel & Das, 2010).

## CONCLUSION

The results of this study suggest that both the PH42 and the I.ROC12 are useful to quantitatively measure aspects of positive health which can be summarised in sum scores in a general population. For both tools, the dimensions and item division differed from the original dialogue tools. This means that for quantitative measurement purposes the dimensions found in our study should be used instead of the dimensions of the dialogue tools. The PH42 is the first choice, however the I.ROC12 is an adequate alternative. For the PH42, further research into item reduction is recommended. For both tools, future research should focus on assessing construct validity (in a general population) in more depth.

## Data Availability

All data from the LISS panel are anonymised and available upon request for researchers and policymakers. For more information see: https://www.lissdata.nl/access-data.

## REFERENCES

Anthony, W. (1993). Recovery from mental illness: The guiding vision of the mental health service system in the 1990s. Psychosocial Rehabilitation Journal, 4, 11–23.

Beckers, T., Koekkoek, B., Hutschemaekers, G., Rudd, B., & Tiemens, B. (2022). Measuring personal recovery in a low-intensity community mental healthcare setting: validation of the Dutch version of the individual recovery outcomes counter (I.ROC). BMC Psychiatry, 22(1), 38. 10.1186/s12888-022-03697-6

Byrne, B. M. (2010). Structural Equational Modeling with AMOS blue book.

Central Commitee on research involving human subjects (CCMO). (n.d.). Questionnaire research.

Davidson, L., & Roe, D. (2007). Recovery from versus recovery in serious mental illness: One strategy for lessening confusion plaguing recovery. Journal of Mental Health, 16(4), 459–470. 10.1080/09638230701482394

de Vet, H. C. W., Terwee, C. B., Mokkink, L. B., & Knol, D. L. (2018). Measurement in Medicine. Cambridge university press: United Kingdom.

Dickens, G. L., Rudd, B., Hallett, N., Ion, R. M., & Hardie, S. M. (2017). Factor validation and Rasch analysis of the individual recovery outcomes counter. Disability and Rehabilitation, 41(1), 74–85. 10.1080/09638288.2017.1375030

Doornenbal, B. M., Vos, R. C., Van Vliet, M., Kiefte-De Jong, J. C., & van den Akker-van Marle, M. E. (2021). Measuring positive health: Concurrent and factorial validity based on a representative Dutch sample. Health and Social Care in the Community, November, 1–9. 10.1111/hsc.13649

Floyd, F. J., & Widaman, K. F. (1995). Factor Analysis in the Development and Refinement of Clinical Assessment Instruments. Psychological Assessment, 7(3), 286–299. 10.1037/1040-3590.7.3.286

Hu, L. T., & Bentler, P. M. (1999). Cutoff criteria for fit indexes in covariance structure analysis: Conventional criteria versus new alternatives. Structural Equation Modeling, 6(1), 1–55. 10.1080/10705519909540118

Huber, M., Knottnerus, J. A., Green, L., Horst, H. v d, Jadad, A. R., Kromhout, D., Leonard, B., Lorig, K., Loureiro, M. I., Meer, J. W. M. v d, Schnabel, P., Smith, R., Weel, C. v, & Smid, H. (2011). How should we define health? British Medical Journal, 343(2), d4163–d4163. 10.1136/bmj.d4163

Huber, M., van Vliet, M., Giezenberg, M., Winkens, B., Heerkens, Y., Dagnelie, P. C., & Knottnerus, J. A. (2016). Towards a patient-centred operationalisation of the new dynamic concept of health. British Medical Journal Open, 6(1), 1–12. 10.1136/bmjopen-201501009110.1136/bmjopen-2015-010091

Institute for positive health. (n.d.). Mijn positieve gezondheid. https://mijnpositievegezondheid.nl/

Ion, R., Monger, B., Hardie, S., Henderson, N., Cumming, J., Hardie, S., & Cumming, J. (2013). A tool to measure progess and Outcome in Recovery. British Journal of Mental Health Nursing, 2, 211–215.

Leamy, M., Bird, V., Boutillier, C. Le, Williams, J., & Slade, M. (2011). Conceptual framework for personal recovery in mental health: systematic review and narrative synthesis. British Journal of Psychiatry, 199(6), 445–452. 10.1192/bjp.bp.110.083733

Monger, B., Hardie, S. M., Ion, R., Cumming, J., & Henderson, N. (2013). The Individual Recovery Outcomes Counter: preliminary validation of a personal recovery measure. The Psychiatrist, 37(7), 221–227. 10.1192/pb.bp.112.041889

Monger, B., Ion, R., Henderson, N., Cumming, J., & Hardie, S. (2012). Outcome measurement in a Scottish mental health charity. Mental Health Today (Brighton, England), April, 24–27.

Prinsen, C. A. C., & Terwee, C. B. (2019). Measuring positive health: for now, a bridge too far. Public Health, 170, 70–77. 10.1016/j.puhe.2019.02.024

Roze, K. C. M., Tijsseling, C., Rudd, B., & Tiemens, B. G. (2020). Measuring Recovery in Deaf, Hard-of-Hearing, and Tinnitus Patients in a Mental Health Care Setting: Validation of the I.ROC. The Journal of Deaf Studies and Deaf Education, 25(2), 178–187. 10.1093/deafed/enz043

Rudd, B. (2018). A holestic psychometric analysis of the individual recovery outcomes counter: balancing user needs in the use of personal outcome measures. Abertay University, School of Social and Health Sciences.

Rudd, B., Karatzias, T., Bradley, A., Fyvie, C., & Hardie, S. (2020). Personally meaningful recovery in people with psychological trauma: Initial validity and reliability of the Individual Recovery Outcomes Counter (I.ROC). International Journal of Mental Health Nursing, 29(3), 387–398. 10.1111/inm.12671

Scherpenzeel, A. C., & Das, M. (2010). “True” Longitudinal and Probability-Based Internet Panels: Evidence From the Netherlands. In Social and Behavioral Research and the Internet: Advances in Applied Methods and Research Strategies. (pp. 77–104). Boca Raton: Taylor & Francis.

Sportel, B. E., Aardema, H., Boonstra, N., Arends, J., Rudd, B., Metz, M. J., Castelein, S., & Pijnenborg, G. H. M. (2023). Measuring recovery in participants with a schizophrenia spectrum disorder: validation of the Individual Recovery Outcomes Counter (I.ROC). BMC Psychiatry, 23(1), 296. 10.1186/s12888-023-04763-3

Tabachnick, B., & Fidell, L. L. S. (2007). Using Multivariate Statistics. Pearson/Allyn & Bacon.

The World Health Organization (WHO). (2006). The World Health Organization (WHO) (Issue April 1948). 10.4324/9780203029732

van Druten, V. P., Bartels, E. A., van de Mheen, D., de Vries, E., Kerckhoffs, A. P. M., & Nahar-van Venrooij, L. M. W. (2022). Concepts of health in different contexts: a scoping review. BMC Health Services Research, 22(1). 10.1186/s12913-022-07702-2

Van Vliet, M., Doornenbal, B. M., Boerema, S., & Van Den Akker-Van Marle, E. M. (2021). Development and psychometric evaluation of a Positive Health measurement scale: A factor analysis study based on a Dutch population. BMJ Open, 11(2), 1–10. 10.1136/bmjopen-2020-040816

Velicer, W. F., & Jackson, D. N. (1990). Component Analysis versus Common Factor Analysis: Some Issues in Selecting an Appropriate Procedure. Multivariate Behavioral Research, 25(1), 1–28.

