## Supplementary tables 1 and 2 for "Measuring positive health using the My Positive Health (MPH) and Individual Recovery Outcomes Counter (I.ROC) dialogue tools: a panel study on measurement properties in a representative general Dutch population"

**Supplementary table 1: items of the my positive health dialogue tool**

| Item number of the dialogue tool | Item |
|---|---|
| BF1 | I feel healthy |
| BF2 | I feel fit |
| BF3 | I don't have complaints or pain |
| BF4 | I sleep well |
| BF5 | I eat well |
| BF6 | I recover quickly after effort. For example after exercise. |
| BF7 | I can move easily. For example, climbing stairs, walking or cycling |
| MW8 | I can remember things well |
| MW9 | I can concentrate well |
| MW10 | I can see, hear, talk, read |
| MW11 | I feel happy |
| MW12 | I accept myself the way I am |
| MW13 | I search for solutions to change difficult situations |
| MW14 | I have control over my life |
| MF15 | I have a meaningful life |
| MF16 | In the morning I'm looking forward to the day |
| MF17 | I have ideals that I would like to achieve |
| MF18 | I have faith in my own future |
| MF19 | I accept life as it comes |
| MF20 | I am grateful for what life offers me |
| MF21 | I want to keep learning all my life |
| QL22 | I am enjoying my life |
| QL23 | I am happy |
| QL24 | I am feeling good |
| QL25 | I experience balance in my life |
| QL26 | I am feeling safe |
| QL27 | I am satisfied with where I live and with whom |
| QL28 | I have enough money to pay my bills |
| SP29 | I have enough contact with other people |
| SP30 | Other people take me seriously |
| SP31 | I have people I can do fun things with |
| SP32 | I have people who support me when needed |
| SP33 | I have the feeling that I 'fit in' in my environment |
| SP34 | I have work or other activities that I find meaningful |
| SP35 | I am interested in what is happening in society |
| DF36 | I can take good care of myself. For example washing, dressing, shopping, cooking |
| DF37 | I know what I can do and what I can't |
| DF38 | I know how to take care of my health |
| DF39 | I can plan well what to do in a day |
| DF40 | I can handle the money I get every month well |
| DF41 | I can work or do voluntary work |
| DF42 | I know how to get help from official authorities if needed |

BF: bodily functions, MW: mental wellbeing, MF: meaningfulness, QL: quality of life, SP: social and societal participation, DF: daily functioning (Huber et al., 2016).

| Supplementary table 2: Items of the individual recovery outcomes counter | |
|---|---|
| Item number of questionnaire | Description |
| HO1 | In the past 3 months… How often have you felt mentally & emotionally healthy, happy and well? |
| HO2 | In the past 3 months… How often have you felt you have the skills you need to look after yourself? |
| HO3 | In the past 3 months… How often have you felt safe and comfortable in and around your home? |
| OP4 | In the past 3 months… How often have you felt physically healthy? |
| OP5 | In the past 3 months… How often would you say you have been active or exercised – on a regular basis? |
| OP6 | In the past 3 months… How often would you say you have felt purposefully occupied? |
| PE7 | In the past 3 months… How often have you felt that you have people/friends/loved ones who can support you if you need it? |
| PE8 | In the past 3 months… How regularly have you taken part in community/group activities? |
| PE9 | In the past 3 months… How often have you felt that you have been able to value and respect yourself? |
| EM10 | In the past 3 months… How often have you felt involved in the decisions that affect your life? |
| EM11 | In the past 3 months… How often have you felt in control and able to manage your life? |
| EM12 | In the past 3 months… How often have you felt hopeful for the future? |

HO: Home, OP: opportunity, PE: people, EM: empowerment (Ion et al., 2013; Monger et al., 2013).

### REFERENCES


Huber, M., van Vliet, M., Giezenberg, M., Winkens, B., Heerkens, Y., Dagnelie, P. C., & Knottnerus, J. A. (2016). Towards a patient-centred operationalisation of the new dynamic concept of health. *British Medical Journal Open*, *6*(1), 1–12. https://doi.org/10.1136/bmjopen-201501009110.1136/bmjopen-2015-010091

Ion, R., Monger, B., Hardie, S., Henderson, N., Cumming, J., Hardie, S., & Cumming, J. (2013). A tool to measure progess and Outcome in Recovery. *British Journal of Mental Health Nursing*, *2*, 211–215.

Monger, B., Hardie, S. M., Ion, R., Cumming, J., & Henderson, N. (2013). The Individual Recovery Outcomes Counter: preliminary validation of a personal recovery measure. *The Psychiatrist*, *37*(7), 221–227. https://doi.org/10.1192/pb.bp.112.041889
