## Supplementary tables 3 to 8. for "Measuring positive health using the My Positive Health (MPH) and Individual Recovery Outcomes Counter (I.ROC) dialogue tools: a panel study on measurement properties in a representative general Dutch population"

**Supplementary table 3: MPH Inter-Item Correlation Matrix of factor Acceptation, meaningfulness and satisfaction with life**

| | QL23 Being happy | QL22 Enjoyment | QL25 Feeling well-balanced | QL24 Feeling good | MF16 Being high-spirited | MW11 Being cheerful | MF15 Having a meaningful life | MF20 Being grateful | MW18 Feeling confident about own future | MF19 Accepting life | MW14 Having control | MW12 Accepting yourself | QL26 Feeling safe |
|---|---|---|---|---|---|---|---|---|---|---|---|---|---|
| QL23 Being happy | 1.000 | 0.893 | 0.811 | 0.844 | 0.784 | 0.815 | 0.769 | 0.756 | 0.746 | 0.664 | 0.699 | 0.652 | 0.646 |
| QL22 Enjoyment | 0.893 | 1.000 | 0.804 | 0.842 | 0.793 | 0.822 | 0.785 | 0.769 | 0.757 | 0.678 | 0.705 | 0.669 | 0.658 |
| QL25 Feeling well-balanced | 0.811 | 0.804 | 1.000 | 0.831 | 0.764 | 0.755 | 0.733 | 0.702 | 0.723 | 0.689 | 0.709 | 0.670 | 0.640 |
| QL24 Feeling good | 0.844 | 0.842 | 0.831 | 1.000 | 0.784 | 0.827 | 0.726 | 0.682 | 0.724 | 0.657 | 0.680 | 0.687 | 0.648 |
| MF16 Being high-spirited | 0.784 | 0.793 | 0.764 | 0.784 | 1.000 | 0.773 | 0.770 | 0.697 | 0.738 | 0.670 | 0.669 | 0.635 | 0.597 |
| MW11 Being cheerful | 0.815 | 0.822 | 0.755 | 0.827 | 0.773 | 1.000 | 0.721 | 0.691 | 0.723 | 0.654 | 0.676 | 0.653 | 0.634 |
| MF15 Having a meaningful life | 0.769 | 0.785 | 0.733 | 0.726 | 0.770 | 0.721 | 1.000 | 0.716 | 0.750 | 0.663 | 0.710 | 0.631 | 0.614 |
| MF20 Being grateful | 0.756 | 0.769 | 0.702 | 0.682 | 0.697 | 0.691 | 0.716 | 1.000 | 0.712 | 0.709 | 0.653 | 0.647 | 0.613 |
| MW18 Feeling confident about own future | 0.746 | 0.757 | 0.723 | 0.724 | 0.738 | 0.723 | 0.750 | 0.712 | 1.000 | 0.697 | 0.715 | 0.640 | 0.623 |
| MF19 Accepting life | 0.664 | 0.678 | 0.689 | 0.657 | 0.670 | 0.654 | 0.663 | 0.709 | 0.697 | 1.000 | 0.664 | 0.667 | 0.596 |
| MW14 Having control | 0.699 | 0.705 | 0.709 | 0.680 | 0.669 | 0.676 | 0.710 | 0.653 | 0.715 | 0.664 | 1.000 | 0.670 | 0.639 |
| MW12 Accepting yourself | 0.652 | 0.669 | 0.670 | 0.687 | 0.635 | 0.653 | 0.631 | 0.647 | 0.640 | 0.667 | 0.670 | 1.000 | 0.574 |
| QL26 Feeling safe | 0.646 | 0.658 | 0.640 | 0.648 | 0.597 | 0.634 | 0.614 | 0.613 | 0.623 | 0.596 | 0.639 | 0.574 | 1.000 |

MW: mental wellbeing, MF: meaningfulness, QL: quality of life (Huber et al., 2016).

**Supplementary table 4: MPH Inter-Item Correlation Matrix between items of factor Physical health and functioning**

| | BF2 Feeling fit | BF7 Exercise | BF1 Feeling healthy | BF6 Physical condition | DF41 Being able to work | BF4 Sleeping pattern | BF5 Eating pattern | BF3 Having complaints or pain |
|---|---|---|---|---|---|---|---|---|
| BF2 Feeling fit | 1.000 | 0.704 | 0.845 | 0.735 | 0.490 | 0.488 | 0.516 | 0.361 |
| BF7 Exercise | 0.704 | 1.000 | 0.682 | 0.735 | 0.548 | 0.395 | 0.435 | 0.312 |
| BF1 Feeling healthy | 0.845 | 0.682 | 1.000 | 0.674 | 0.518 | 0.488 | 0.476 | 0.348 |
| BF6 Physical condition | 0.735 | 0.735 | 0.674 | 1.000 | 0.477 | 0.462 | 0.537 | 0.313 |
| DF41 Being able to work | 0.490 | 0.548 | 0.518 | 0.477 | 1.000 | 0.283 | 0.294 | 0.184 |
| BF4 Sleeping pattern | 0.488 | 0.395 | 0.488 | 0.462 | 0.283 | 1.000 | 0.529 | 0.262 |
| BF5 Eating pattern | 0.516 | 0.435 | 0.476 | 0.537 | 0.294 | 0.529 | 1.000 | 0.256 |
| BF3 Having complaints or pain | 0.361 | 0.312 | 0.348 | 0.313 | 0.184 | 0.262 | 0.256 | 1.000 |

BF: bodily functions, DF: daily functioning (Huber et al., 2016).

**Supplementary table 5: MPH Inter-Item Correlation Matrix between items of factor Self-management**

| | DF40 Managing money | DF37 Knowing your limitations | DF38 Knowledge of health | DF36 Looking after yourself | DF39 Managing time | QL28 Having enough money | DF42 Asking for help |
|---|---|---|---|---|---|---|---|
| DF40 Managing money | 1.000 | 0.569 | 0.570 | 0.503 | 0.554 | 0.690 | 0.443 |
| DF37 Knowing your limitations | 0.569 | 1.000 | 0.779 | 0.656 | 0.628 | 0.403 | 0.393 |
| DF38 Knowledge of health | 0.570 | 0.779 | 1.000 | 0.666 | 0.602 | 0.413 | 0.404 |
| DF36 Looking after yourself | 0.503 | 0.656 | 0.666 | 1.000 | 0.501 | 0.359 | 0.305 |
| DF39 Managing time | 0.554 | 0.628 | 0.602 | 0.501 | 1.000 | 0.388 | 0.417 |
| QL28 Having enough money | 0.690 | 0.403 | 0.413 | 0.359 | 0.388 | 1.000 | 0.379 |
| DF42 Asking for help | 0.443 | 0.393 | 0.404 | 0.305 | 0.417 | 0.379 | 1.000 |

QL: quality of life, DF: daily functioning (Huber et al., 2016).

**Supplementary table 6: MPH Inter-Item Correlation Matrix between items of factor Social network and societal roles**

| | SP31 Doing fun things together | SP32 Having the support of others | SP33 Belonging | SP29 Social contacts | SP30 Being taken seriously | SP34 Doing meaningful things | QL27 Living conditions | SP35 Being interested in society |
|---|---|---|---|---|---|---|---|---|
| SP31 Doing fun things together | 1.000 | 0.822 | 0.774 | 0.734 | 0.694 | 0.598 | 0.538 | 0.492 |
| SP32 Having the support of others | 0.822 | 1.000 | 0.779 | 0.695 | 0.678 | 0.557 | 0.582 | 0.467 |
| SP33 Belonging | 0.774 | 0.779 | 1.000 | 0.743 | 0.711 | 0.621 | 0.569 | 0.523 |
| SP29 Social contacts | 0.734 | 0.695 | 0.743 | 1.000 | 0.724 | 0.607 | 0.509 | 0.481 |
| SP30 Being taken seriously | 0.694 | 0.678 | 0.711 | 0.724 | 1.000 | 0.561 | 0.533 | 0.540 |
| SP34 Doing meaningful things | 0.598 | 0.557 | 0.621 | 0.607 | 0.561 | 1.000 | 0.485 | 0.521 |
| QL27 Living conditions | 0.538 | 0.582 | 0.569 | 0.509 | 0.533 | 0.485 | 1.000 | 0.391 |
| SP35 Being interested in society | 0.492 | 0.467 | 0.523 | 0.481 | 0.540 | 0.521 | 0.391 | 1.000 |

QL: quality of life, SP: social and societal participation (Huber et al., 2016).

**Supplementary table 7: MPH Inter-Item Correlation Matrix between items of factor Personal development**

| | MF21 Continue learning | MF17 Wanting to achieve ideals | MW13 Being able to handle changes |
|---|---|---|---|
| MF21 Continue learning | 1.000 | 0.534 | 0.483 |
| MF17 Wanting to achieve ideals | 0.534 | 1.000 | 0.449 |
| MW13 Being able to handle changes | 0.483 | 0.449 | 1.000 |

MW: mental wellbeing, MF: meaningfulness (Huber et al., 2016).

**Supplementary table 8: MPH Inter-Item Correlation Matrix between items of factor Cognition**

| | MW8 Being able to remember things | MW9 Being able to concentrate | MW10 Being able to communicate |
|---|---|---|---|
| MW8 Being able to remember things | 1.000 | 0.768 | 0.477 |
| MW9 Being able to concentrate | 0.768 | 1.000 | 0.452 |
| MW10 Being able to communicate | 0.477 | 0.452 | 1.000 |

MW: mental wellbeing (Huber et al., 2016).

### REFERENCES

Huber, M., van Vliet, M., Giezenberg, M., Winkens, B., Heerkens, Y., Dagnelie, P. C., & Knottnerus, J. A. (2016). Towards a patient-centred operationalisation of the new dynamic concept of health. *British Medical Journal Open*, *6*(1), 1–12. https://doi.org/10.1136/bmjopen-201501009110.1136/bmjopen-2015-010091
