## Supplementary tables 9 and 10 for "Measuring positive health using the My Positive Health (MPH) and Individual Recovery Outcomes Counter (I.ROC) dialogue tools: a panel study on measurement properties in a representative general Dutch population"

**Supplementary table 9: I.ROC Inter-Item Correlation Matrix between items of factor Wellbeing, control, network and meaningfulness**

| | EM12 Self-management | EM10 Participation and control | PE9 Valuing myself | EM11 Hope for the future | PE7 Personal network | OP6 Purpose and direction | HO1 Mental health | PE8 Social network |
|---|---|---|---|---|---|---|---|---|
| EM12 Self-management | 1.000 | 0.553 | 0.584 | 0.614 | 0.487 | 0.534 | 0.582 | 0.390 |
| EM10 Participation and control | 0.553 | 1.000 | 0.602 | 0.666 | 0.488 | 0.479 | 0.470 | 0.350 |
| PE9 Valuing myself | 0.584 | 0.602 | 1.000 | 0.563 | 0.455 | 0.537 | 0.554 | 0.341 |
| EM11 Hope for the future | 0.614 | 0.666 | 0.563 | 1.000 | 0.439 | 0.442 | 0.554 | 0.301 |
| PE7 Personal network | 0.487 | 0.488 | 0.455 | 0.439 | 1.000 | 0.437 | 0.436 | 0.388 |
| OP6 Purpose and direction | 0.534 | 0.479 | 0.537 | 0.442 | 0.437 | 1.000 | 0.499 | 0.356 |
| HO1 Mental health | 0.582 | 0.470 | 0.554 | 0.554 | 0.436 | 0.499 | 1.000 | 0.295 |
| PE8 Social network | 0.390 | 0.350 | 0.341 | 0.301 | 0.388 | 0.356 | 0.295 | 1.000 |

HO: Home, OP: opportunity, PE: people, EM: empowerment (Ion et al., 2013; Monger et al., 2013).

**Supplementary table 10: I.ROC Inter-Item Correlation Matrix between items of factor Health, safety and abilities**

| | OP5 Exercise and activity | HO2 Life skills | OP4 Physical health | HO3 Safety and comfort |
|---|---|---|---|---|
| OP5 Exercise and activity | 1.000 | 0.369 | 0.527 | 0.287 |
| HO2 Life skills | 0.369 | 1.000 | 0.458 | 0.485 |
| OP4 Physical health | 0.527 | 0.458 | 1.000 | 0.399 |
| HO3 Safety and comfort | 0.287 | 0.485 | 0.399 | 1.000 |

HO: Home, OP: opportunity (Ion et al., 2013; Monger et al., 2013).

### REFERENCES

Ion, R., Monger, B., Hardie, S., Henderson, N., Cumming, J., Hardie, S., & Cumming, J. (2013). A tool to measure progess and Outcome in Recovery. *British Journal of Mental Health Nursing*, *2*, 211–215.

Monger, B., Hardie, S. M., Ion, R., Cumming, J., & Henderson, N. (2013). The Individual Recovery Outcomes Counter: preliminary validation of a personal recovery measure. *The Psychiatrist*, *37*(7), 221–227. https://doi.org/10.1192/pb.bp.112.041889
